# Effectiveness and Adverse Event Profiles of Catheter Ablation in Persistent Atrial Fibrillation: A Meta–Analysis of Randomized and Single–Arm Clinical Trials

**DOI:** 10.64898/2026.05.27.26354285

**Authors:** Harizavi Arshan, Yan Chai, Jiayuan Wang, Tiffany Tan

**Affiliations:** Johnson and Johnson, Biosense Webster Inc, Cardiac Electrophysiology, Clinical Data Science, Irvine, California, United States; Medical Informatics, University of California Los Angeles, Los Angeles, California, United States

**Keywords:** persistent atrial fibrillation, catheter ablation, pulmonary vein isolation, radiofrequency ablation, pulsed field ablation, arrhythmia recurrence, procedural complications, ablation adverse events, stroke, meta-analysis, clinical effectiveness, procedural efficiency

## Abstract

**BACKGROUND:** Catheter ablation is an established rhythm-control strategy for atrial fibrillation, but outcomes in persistent atrial fibrillation (PsAF) remain heterogeneous across evolving strategies and energy modalities. An updated synthesis is needed to define current effectiveness and adverse-event profiles in the modern ablation era.

**METHODS:** We conducted a systematic review and meta-analysis of prospective clinical trials of catheter ablation for PsAF published from 2010 through December 2025. We included randomized and nonrandomized prospective interventional studies reporting effectiveness and adverse events, and pooled outcomes using random-effects models. Prespecified subgroup analyses evaluated ablation strategy (pulmonary vein isolation [PVI] vs PVI with adjunctive lesion sets [PVI+]), ablation modality (radiofrequency [RF], cryoballoon [CRYO], and pulsed field [PF]), and endpoint definition (recurrence-only vs composite measures).

**RESULTS:** Thirty-two studies (9,194 patients) met inclusion criteria; 28 (7,948 patients) contributed to effectiveness analyses. The pooled 12-month arrhythmia-free proportion was 0.65 (95% CI, 0.61–0.68), with substantial heterogeneity. Effectiveness was numerically higher with PVI+ than PVI-only (0.66 [0.60–0.72] vs 0.63 [0.59–0.67]), similar for PF (0.65 [0.57–0.72]) and RF (0.65 [0.61–0.69]), and slightly lower for CRYO (0.64 [0.54–0.74]). Recurrence-only endpoints yielded higher effectiveness than composite endpoints (0.67 [0.63–0.71] vs 0.60 [0.55–0.64]). Safety analyses included 32 studies (9,002 patients). Adverse events were low but heterogeneous (0%–14.56%); pooled vascular access and pericardial complication incidences were each 1%, while thromboembolic events, accessory organ injury, and mortality were rare (pooled 0%). PF ablation showed numerically lower overall complication incidences than RF and CRYO.

**CONCLUSION:** In contemporary trials, catheter ablation for PsAF shows moderate effectiveness and low overall adverse-event risk. Adjunctive strategies and PF ablation are promising, but no approach is consistently superior. These findings support tailored, patient-specific ablation selection in PsAF.

Systematic Review Registration: PROSPERO, https://www.crd.york.ac.uk/prospero/, identifier 1322250.

**CLINICAL RELEVANCE:** *What Is New?:*

- This meta-analysis provides an updated synthesis of catheter ablation outcomes in persistent atrial fibrillation, including contemporary technologies and strategies such as pulsed field ablation and adjunctive lesion sets beyond pulmonary vein isolation.
- Contemporary trials demonstrated moderate effectiveness, with an overall 12-month arrhythmia-free proportion of approximately 65% and substantial heterogeneity across studies.
- Adverse events were uncommon, with vascular access and pericardial complications each occurring in about 1% of patients and other major complications remaining rare.

*What Are the Clinical Implications?:*

- Catheter ablation is a safe, effective rhythm-control option for persistent atrial fibrillation and should be considered in patient-centered care.
- Clinicians should interpret outcomes within the context of significant variability across studies and tailor procedural strategies based on individual patient characteristics and operator expertise.
- Further randomized studies with standardized endpoints and monitoring are needed to define optimal approaches and improve long-term outcomes.

## INTRODUCTION

Atrial fibrillation (AF) is the most common sustained arrhythmia worldwide, affecting over 50 million people and increasing the risk of stroke, heart failure, myocardial infarction, dementia, and death [1]. Its prevalence continues to rise; an estimated 1 in 3 to 5 adults over age 45 will develop AF, and U.S. prevalence is projected to roughly double over the next decade [2]. AF is classified as paroxysmal or persistent based on episode duration and spontaneous termination. Compared with paroxysmal AF (PAF), persistent AF (PsAF) reflects more advanced atrial structural and electrical remodeling and is associated with greater arrhythmia burden and worse outcomes, including heart failure and all-cause mortality [3].

Over the past three decades, catheter ablation (CA) has become an effective rhythm-control strategy for AF, and the 2023 ACC/AHA/ACCP/HRS Guideline for the Diagnosis and Management of Atrial Fibrillation states that first-line CA may be reasonable for selected patients with symptomatic PsAF [4]. This shift reflects a rapidly evolving treatment landscape, including newer energy sources such as pulsed field ablation and expanded strategies beyond pulmonary vein isolation (PVI), such as guided lesion creation, substrate modification (SM), and posterior wall isolation (PWI) [14, 15, 27].

As ablation becomes more established for PsAF, its contemporary performance across evolving strategies and technologies should be reassessed. This meta-analysis provides an updated overview of clinical trials evaluating the effectiveness and adverse event profiles of CA for PsAF, with prespecified analyses by ablation strategy and energy modality.

## METHODS

We performed a systematic review and meta-analysis of prospective single-arm trials and randomized controlled trials (RCTs) assessing the effectiveness and adverse events of CA in PsAF. The review followed PRISMA [5] and AMSTAR [6] guidelines. The protocol was prespecified and prospectively registered with PROSPERO (ID: 1322250), with only minor subsequent amendments.

## SEARCH STRATEGY

A comprehensive literature search was conducted in PubMed, EMBASE, the Cochrane Library, and Google Scholar, covering all publications from January 2010 through December 2025. The search strategy incorporated both controlled vocabulary and free-text keywords related to PsAF and CA, including “persistent atrial fibrillation,” “catheter ablation,” “pulmonary vein isolation,” “adjunctive lesion sets,” “posterior wall isolation,” and “linear ablation,” as well as specific energy modalities such as “radiofrequency,” “cryoballoon,” and “pulsed field.” Boolean operators (“AND” and “OR”) were applied to refine and combine search terms.

## STUDY SELECTION

The search was limited to studies from January 2010 onward to capture current ablation technologies and mapping systems. Eligible studies were prospective single-arm trials or RCTs of CA for PsAF reporting 12-month arrhythmia recurrence after a predefined blanking period, procedural complications, or adverse events. Two reviewers (YC and AH) independently performed study identification, screening, and eligibility assessment per PRISMA, resolving disagreements by consensus.

## DATA EXTRACTION

Data were mapped to predefined categories by two reviewers (YC, AH), with discrepancies resolved by a third (JW).

### Effectiveness Endpoint

We used the primary effectiveness endpoint defined in each eligible study as the effectiveness outcome. Most studies defined trial success as freedom from arrhythmia recurrence alone, while some studies used composite endpoints (e.g., freedom from recurrence, repeat ablation, and antiarrhythmic drug [AAD] use). Only studies with a standard 3-month blanking period after the index procedure were included. Primary effectiveness endpoints for each study are listed in Table S1.

When studies reported primary effectiveness endpoint as a proportion, the corresponding number of successes was extracted directly when available. If only Kaplan–Meier estimates were provided, the number of successes was calculated based on the reported survival rates and sample sizes.

### Adverse Event Categorization

The frequency of each reported adverse event within 12 months of the index procedure was extracted and classified into five predefined categories: (1) vascular access complications (hematoma, transfusion-requiring bleeding, pseudoaneurysm, arteriovenous fistula); (2) pericardial complications (pericardial effusion, cardiac tamponade); (3) thromboembolic events (stroke, transient ischemic attack); (4) accessory organ injuries (atrioesophageal fistula, phrenic nerve injury, pulmonary vein stenosis); and (5) all-cause mortality. Categories were chosen for clinical relevance, consistency across contemporary trials, and alignment with consensus-defined major complications. Adverse event definitions and classification were based on U.S. Food and Drug Administration guidance for AF ablation trials (Docket No. FDA-2009-D-0395) [7], as well as the latest international consensus statement on CA of AF [8].

To improve comparability across studies, events were grouped as reported and adjudicated in the original publications and were not reclassified by inferred severity, timing, or procedural attribution. Events were included regardless of timing, from peri-procedural through follow-up to 12 months, to capture total adverse event burden. When trials reported multiple eligible treatment arms, events were pooled across arms. Analyses focused on total event counts and cumulative incidence within each category across studies.

### Subgroup Categorization

#### Ablation Strategy

Ablation strategies were categorized as PVI-only or PVI with adjunctive lesion sets (PVI+), including PWI and other SM approaches. Any trial arm employing additional lesions beyond PVI was classified as PVI+, regardless of specific lesion combinations. In cases where the ablation strategy was not explicitly detailed in the trials, procedural data were reviewed to identify any reports of PVI+. Data were extracted according to ablation strategy when separate results were available; otherwise, studies were assigned to the combined PVI/PVI+ group if PVI+ had been applied but not reported separately.

#### Catheter Energy Type

Studies included pulsed field (PF), cryoballoon (CRYO), and radiofrequency (RF) ablation arms. Data were extracted by energy modality if reported separately.

#### PsAF Population

Some trials included patients with long-standing persistent atrial fibrillation (LsPAF). Studies that allowed LsPAF participants but did not provide separate results for PsAF and LsPAF were included in the analysis to make use of all available evidence. If reported separately, results specific to LsPAF populations were excluded from effectiveness analysis. Studies with unclear PsAF or LsPAF inclusion criteria or unspecified AF type were noted in Table S1.

Studies were evaluated individually and could contribute to multiple analytic areas if justified (e.g., multi-arm trials reporting multiple ablation strategies or endpoint types). Not all trials provided data for every domain due to varying blanking periods, follow-up durations, or endpoint reporting. Only studies meeting prespecified criteria were included in each analysis group. An overview of trial inclusion across analytic domains is provided in Table S1.

## RISK OF BIAS ASSESSMENT

To ascertain the validity of included RCTs, two reviewers (YC and AH) independently assessed the risk of bias (RoB) using the method in the Cochrane Collaboration Handbook for Systematic Reviews of Interventions. RoB was evaluated as high (red), low (green), and unclear (yellow) in each RCT across prespecified domains: randomization process, deviations from intended interventions, missing outcome data, measurement of outcomes, and selection of reported results.

## STATISTICAL ANALYSIS

Analyses were performed in STATA (version 17.0, StataCorp, College Station, TX, USA). Random-effects models were used to address heterogeneity across PsAF ablation studies, with fixed-effects models as sensitivity analyses. Effectiveness and adverse events were summarized as pooled proportions with 95% CIs. No comparative effect measures (e.g., odds ratios or hazard ratios) were calculated. Publication bias was assessed visually using a funnel plot.

## SUBGROUP ANALYSES

We conducted subgroup analyses based on pre-specified categories for both effectiveness and adverse event outcomes. Subgroups were defined according to:

1. Ablation strategy: PVI-only vs. PVI+
2. Catheter energy modality: PF vs. RF vs. CRYO ablation

In addition, a separate subgroup analysis was performed to evaluate effectiveness by primary endpoint definition: arrhythmia recurrence alone versus composite measures (e.g., recurrence, repeat ablation, cardioversion, or AAD use). This analysis evaluated whether pooled effectiveness varied by endpoint definition and helped contextualize differences in reported success rates across studies.

All subgroup analyses were descriptive, using pooled proportions rather than comparative effect measures; observed differences should not be interpreted as evidence of superiority.

## RESULTS

### LITERATURE SEARCH

Database searches identified 1,961 potentially relevant AF ablation studies, including PAF and PsAF treated with PF, RF, or CRYO. After title and abstract screening, 1,874 were excluded as irrelevant, leaving 87 for full-text review. After removing duplicate datasets, reviews, letters, and editorials (n=16), 71 studies were assessed for eligibility; 39 were excluded for ineligible AF subtypes without PsAF stratification, noneligible study designs, nonstandard blanking periods, missing outcomes of interest, procedural-only reporting, or non-AF or non-ablation populations. Ultimately, 32 studies met inclusion criteria and were included in the final meta-analysis (Figure 1).

**Figure 1.**
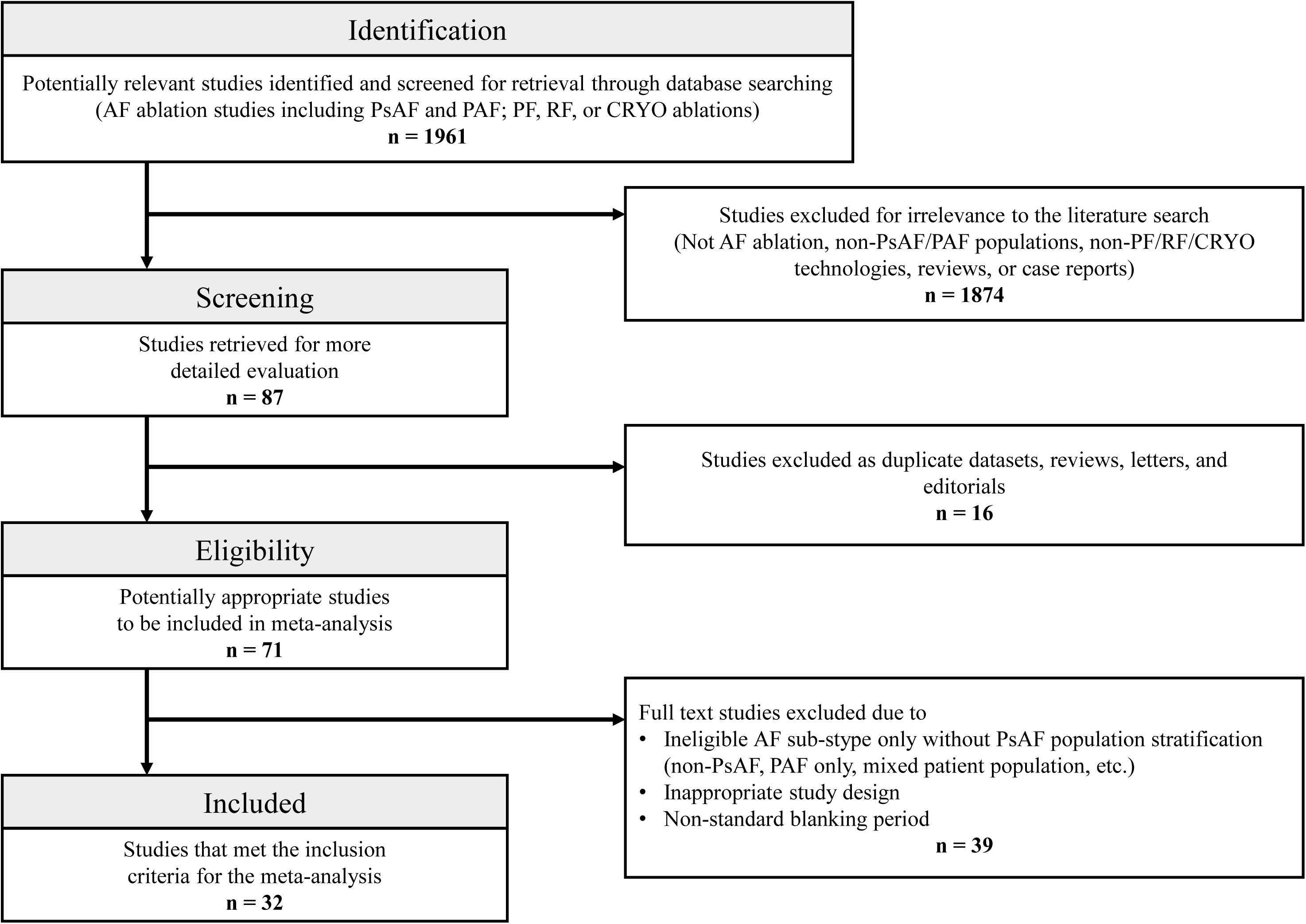
Flow diagram showing study identification, screening, eligibility review, and final inclusion for studies evaluating CA in PsAF.

### STUDY CHARACTERISTICS

We identified 32 studies including 9,194 patients with PsAF undergoing CA [9–40]: 24 RCTs [9–12, 15, 17, 20–22, 25, 27–38, 40, 41] and 8 prospective single-arm trials [13, 14, 16, 18, 19, 23, 24, 26, 39]. Two studies [14, 23] enrolled mixed populations of PAF and PsAF. For trials comparing RF, CRYO, or PF ablation, outcomes from each treatment arm were included in their respective modality groups, but each trial was counted once at the study level, with PsAF patients from both arms contributing to the overall pooled population.

RF was the most common modality, followed by PF and CRYO, with a smaller number of studies evaluating dual-energy approaches combining PF and RF. Ablation strategies included PVI-only and PVI+ approaches, such as PWI, linear lesions, SM, vein of Marshall ethanol infusion (EIVOM), electrogram-guided ablation, or AI-guided targeting. Study designs, sample sizes, strategies, effectiveness, and adverse event incidence are summarized in Table 1; additional study details are provided in Table S1.

**Table 1.** Characteristics included clinical trials evaluating catheter ablation for persistent atrial fibrillation. PsAF = persistent atrial fibrillation; PVI = pulmonary vein isolation; PVI+ = pulmonary vein isolation with adjunct lesion sets; [1] Study Design: Pros = prospective; SA = single-arm; RCT = randomized controlled trial; NI = non-inferiority; IDE = investigational device exemption; FIH = first-in-human; [2] Region reflects primary enrollment geography based on published trial site locations; [3] Ablation Strategy: PWI = posterior wall isolation; SM = substrate modification; CFAE = complex fractionated atrial electrograms; Lines = linear lesions; EIVOM = vein of Marshall ethanol infusion; Fib = MRI-guided fibrosis ablation; FIRM = focal impulse and rotor modulation; AI-PVI+ = ablation index–guided PVI with adjunct lesions; * Sphere Per-AF = mixed-energy study (pulsed field + radiofrequency), values reported as PF/RF arms; † CONVERGE AF = hybrid epicardial arm excluded, endocardial catheter ablation arm only; ‡ PRECEPT = primary effectiveness endpoint reported at 15-month follow-up; ** EXCAPE-AF = mixed-energy study (radiofrequency/cryoballoon), values reported as RF-PVI/CRYO-PVI/CRYO-PVI+ arms; NR = not reported or not eligible; trials were included if ≥1 study arm met prespecified eligibility criteria.

| Trial (Year) | Design [1] | Region [2] | PsAF (N) | Strategy [3] | 12-Mo Effectiveness (%) | Adverse Event Incidence (%) |
| --- | --- | --- | --- | --- | --- | --- |
| **Pulsed Field Ablation** |  |  |  |  |  |  |
| ADVANTAGE-AF (2025) | Pros, SA (IDE) | Global | 276 | PVI+ | 63.9 | 2.69 |
| ADVANTAGE-AF Phase II (2025) | Pros, SA | Global | 255 | PVI+PWI | 73.5 | 3.14 |
| PULSED AF (2023) | Pros, SA | Global | 150 | PVI | 58.9 | 0.67 |
| Sphere Per-AF (2024)* | RCT NI 1:1 | Global | 219 / 213 | PVI+Lines | 73.8* / 65.8 | 0.94* / 0.96 |
| VOLT CE (2025) | Pros, FIH | Europe | 43 | PVI | 58.1 | 2.33 |
| **Radiofrequency Ablation** |  |  |  |  |  |  |
| CAPLA (2023) | RCT 1:1 | Australia / Canada / UK | 168 / 170 | PVI / PVI+PWI | 53.6 / 52.4 | 2.38 / 2.94 |
| CONVERGE AF (2020)† | RCT 2:1 | US | 51 | PVI+ | 50 | 0 |
| CRRF-PeAF (2025) | RCT 1:1 | Asia | 250 | PVI / PVI ± Adj | 74.7 / 80.0 | 2.00 |
| DECAAF II (2022) | RCT 1:1 | Europe / Australia / US | 422 / 421 | PVI / PVI+Fib | 53.9 / 57.0 | 3.72 / 1.17 |
| EARNEST-PVI (2021) | RCT 1:1 | Asia | 254 / 258 | PVI / PVI+ | 71.5 / 78.2 | 1.61 / 3.23 |
| ERASE-AF (2022) | RCT 1:1 | Germany | 163 / 161 | PVI / PVI+SM | 50.0 / 64.7 | 1.84 / 3.73 |
| IRRIGATE RF (2017) | RCT 1:1 | Europe | 77 | PVI | 71.2 | 4.55 |
| MAP-AF (2024) | RCT 1:1 | Asia | 103 / 103 | PVI / PVI+MAP | NR | 0 / 2.00 |
| MARSHALL-PLAN (2025) | RCT 1:1 | Europe | 59 / 59 | PVI / PVI+EIVOM | 66.1 / 86.4 | 3.39 / 13.56 |
| NO-PERSAF (2022) | RCT 1:1 | Asia | 49 | PVI | 61.2 | 4.08 |
| PeAF by LAWt (2023) | RCT 1:1 | Asia | 78 / 78 | PVI / PVI+LAWT | 77.6 / 81.3 | 2.56 / 6.40 |
| PERSIST-END (2022) | Pros, SA (IDE) | US / Australia | 224 | PVI ± Adj | NR | 3.14 |
| POBI-AF (2019) | RCT 1:1 | Asia | 107 / 106 | PVI / PVI+PWI | 79.0 / 78.4 | 5.71 / 3.92 |
| PRECEPT (2020)‡ | Pros, SA (IDE) | Global | 381 | PVI ± Adj | NR | 4.07 |
| PROMPT-AF (2024) | RCT 1:1 | China | 250 / 248 | PVI / PVI+EIVOM | 61.5 / 70.7 | 5.69 / 1.61 |
| REAFFIRM (2025) | RCT 1:1 | Europe / US | 171 / 179 | FIRM-PVI / FIRM-PVI+ | 71.9 / 64.0 | 7.14 |
| RF vs CRYO PsAF (2023) | RCT 2:1 | Europe | 133 | PVI | 73.7 | 0.75 |
| SARA Study (2014) | RCT 2:1 | Europe | 98 | PVI | 70.4 | 8.16 |
| SCAR-AF (2025) | RCT 1:1 | Europe | 72 / 76 | PVI / PVI+LVA | NR | 2.78 / 5.26 |
| STAR-AF II (2015) | RCT 1:4:4 | Global | 67 / 263 / 259 | PVI / PVI+CFAE / PVI+Lines | 59.0 / 48.8 / 45.9 | 4.69 / 2.76 / 4.40 |
| SUPPRESS-AF (2025) | RCT 1:1 | Asia | 172 / 170 | PVI / PVI+LVA | 50.3 / 61.2 | 2.91 / 5.29 |
| TAILORED-AF (2025) | RCT 1:1 | Global | 188 / 186 | PVI / AI-PVI+ | 70.1 / 87.8 | 3.83 / 6.95 |
| Three-Guided Ablation (2023) | RCT 1:1:1 | China | 150 / 150 / 150 | ANAT / EGM / EXT | 54.0 / 64.0 / 72.0 | 2.67 / 2.67 / 3.33 |
| VENUS (2020) | RCT 2:1 | US | 158 / 185 | PVI / PVI+EIVOM | 38.0 / 49.2 | 14.56 / 13.51 |
| **Cryoballoon Ablation** |  |  |  |  |  |  |
| CRRF-PeAF (2025) | RCT 1:1 | Asia | 249 | PVI / PVI ± Adj | 73.9 / 84.1 | 1.60 |
| CRYO4PERSISTENT AF (2018) | Pros, SA | Europe | 130 | PVI | 67.3 | 2.97 |
| EXCAPE-AF (2025)** | RCT 1:1:1 | South Korea | 94 / 98 / 96 | RF-PVI / CRYO-PVI / CRYO-PVI+ | 51.1 / 49.0 / 45.8 | 1.06 / 0 / 0 |
| NO-PERSAF (2022) | RCT 1:1 | Asia | 52 | PVI | 69.2 | 0 |
| RF vs CRYO PsAF (2023) | RCT 2:1 | Europe | 66 | PVI | 72.7 | 0 |
| STOP Persistent AF (2020) | Pros, SA | Global | 186 | PVI | 50.3 | 4.24 |

### PRIMARY EFFECTIVENESS ENDPOINT ANALYSIS

A total of 28 studies, comprising approximately 7,948 patients with PsAF, contributed data to the primary effectiveness endpoint analysis. Two studies were excluded because they used non-comparable blanking periods of six months rather than the standard three months, and two additional studies were excluded because they did not report a 12-month effectiveness outcome. Follow-up duration was predominantly 12 months, although several studies reported additional time points extending to 15 or 18 months. As summarized in Table S1, monitoring intensity ranged from intermittent electrocardiograms (ECGs) and short-duration Holter recordings to continuous surveillance with implantable cardiac monitors (ICMs) or frequent smartphone-based ECG recordings.

Reported proportion of freedom from atrial arrhythmia at 12 months varied substantially across studies, ranging from approximately 38% to 88%, reflecting differences in study design, energy source, ablation strategy, monitoring intensity, and patient population (Table 1, Table S1).

Using a random-effects model, CA was associated with a pooled estimate of arrhythmia-free proportion at 12 months of 0.65 (95% CI, 0.61–0.68). Between-study heterogeneity was high (I² = 90.3%) as shown in Figure 2.

**Figure 2.**
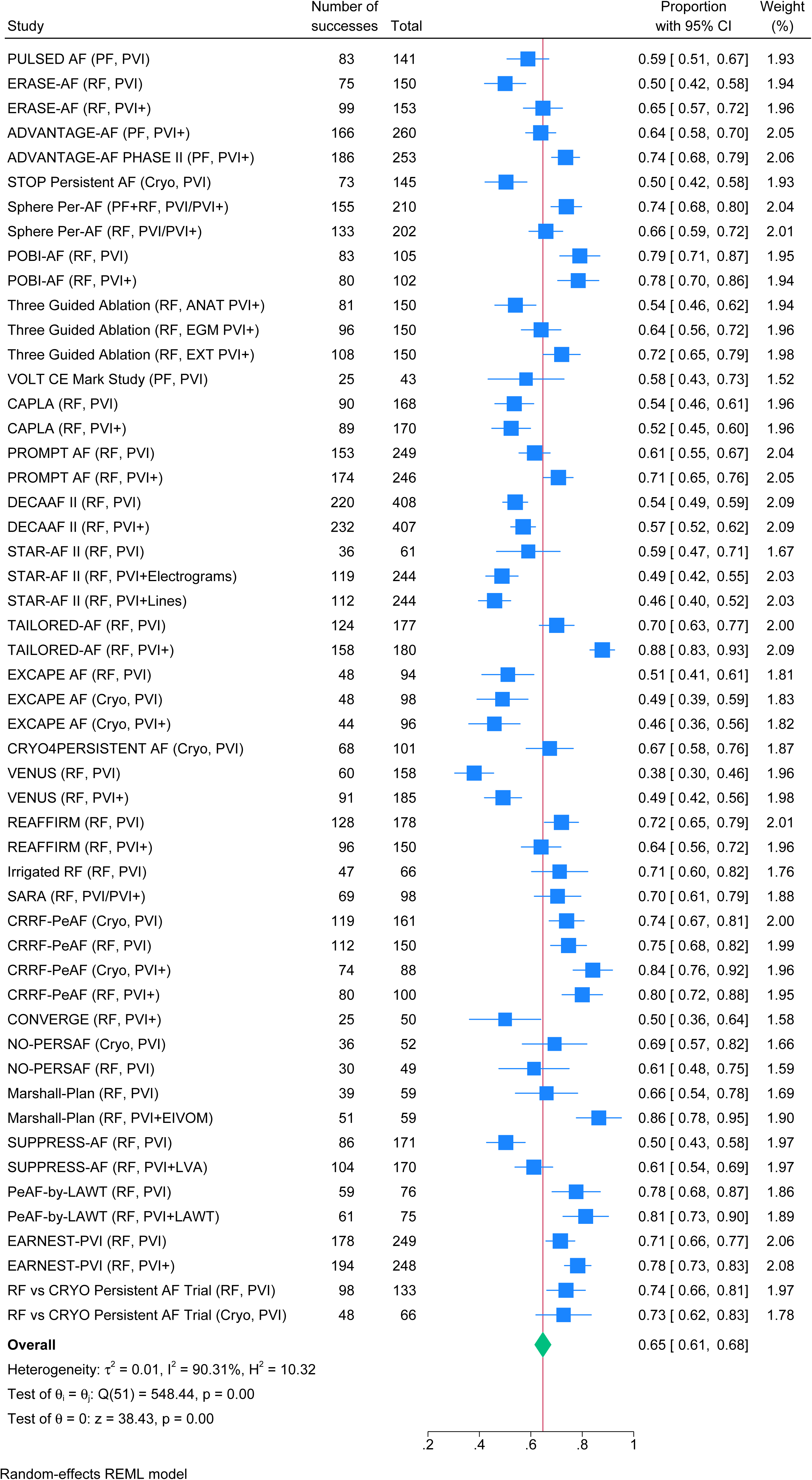
Overall 12-month primary effectiveness following catheter ablation for persistent atrial fibrillation. CI = confidence interval; proportions represent 12-month arrhythmia-free survival or composite primary effectiveness endpoint, as defined by each study; squares indicate study estimates (size proportional to weight) with 95% CIs; diamond indicates pooled estimate; random-effects REML model used; heterogeneity assessed by τ², I², and H².

#### Effectiveness By Ablation Strategy: PVI vs. PVI+

PVI-only and PVI+ were commonly employed strategies across the included trials. Of the 28 studies, 22 contributed data to the PVI-only group, while 19 contributed data to the PVI+ group for effectiveness outcomes. Adjunctive strategies included PWI, linear lesions, complex fractionated atrial electrogram (CFAE) ablation, EIVOM ablation, fibrosis-guided ablation, low-voltage-area ablation, wall-thickness-guided ablation, and other mapping- or algorithm-guided lesion sets.

Using a random-effects model, the pooled 12-month arrhythmia-free proportion was 0.63 (95% CI, 0.59–0.67) for PVI-only, 0.66 (95% CI, 0.60–0.72) for PVI+, and 0.70 (95% CI, 0.65– 0.75) for the few studies with mixed PVI-only and PVI+ populations (Figure S1).

#### Effectiveness By Catheter Energy: RF, CRYO, PF

Among the 28 studies analyzed, 4 provided data for the PF ablation group, 22 for the RF ablation group, and 6 for the CRYO ablation group. The years in which these trials were conducted ranged from 2023 to 2025 for PF ablation, 2014 to 2025 for RF ablation, and 2018 to 2025 for CRYO ablation.

Using a random-effects model, the pooled 12-month arrhythmia-free proportion was 0.64 (95% CI, 0.54–0.74) for CRYO, 0.65 (95% CI, 0.57–0.72) for PF, and 0.65 (95% CI, 0.61–0.69) for RF. The single study [17] with a combined PF+RF approach reported a 12-month arrhythmia-free proportion of 0.74 (95% CI, 0.68–0.80) (Figure S2).

#### Effectiveness By Endpoint Type: Recurrence-Only vs. Composite Measures

Among the 28 studies, 18 used arrhythmia recurrence as their primary effectiveness endpoint, while 10 used composite measures. Freedom from recurrent atrial arrhythmia following CA for PsAF was defined similarly across studies, most commonly as the absence of AF, atrial flutter, or atrial tachycardia lasting ≥30 seconds after a predefined blanking period. Composite effectiveness endpoints frequently incorporated additional failure criteria, including repeat ablation, cardioversion, or escalation of Class I or III AAD therapy (Table 1, Table S1).

Using a random-effects model, the pooled 12-month arrhythmia-free proportion was 0.67 (95% CI, 0.63–0.71) for recurrence-only endpoints and 0.60 (95% CI, 0.55–0.64) for composite endpoints (Figure S3). These findings suggest that endpoint definition influences observed effectiveness outcomes, although differences in pooled estimates were modest (∼5%) and 95% CIs showed substantial overlap.

### ADVERSE EVENT ANALYSIS

A total of 32 unique studies, comprising approximately 9,002 patients with PsAF, contributed data to the adverse-event analysis (Table S1). The included studies comprised randomized and single-arm prospective investigations evaluating RF, CRYO, PF ablation, and hybrid ablation strategies (Table S1).

After systematically abstracting and harmonizing reported adverse events across studies, we mapped events to standardized categories to accommodate variations in adverse event definitions and reporting frameworks (Table S2). Event-capture windows varied by study, ranging from intraprocedural or ≤7-day reporting to 30-day assessments and extended follow-up up to 12 months (Table S1). Frequently reported adverse events included bleeding and vascular complications, cardiac perforation or tamponade, stroke or transient ischemic attack, phrenic nerve injury, esophageal injury, pulmonary vein stenosis, pericardial syndromes, heart block requiring permanent pacemaker implantation, heart failure, and all-cause death, with several studies employing independent clinical event adjudication (Table S2).

Overall, the pooled incidence of adverse events was low but heterogeneous across studies, with reported incidences ranging from 0% to approximately 14.56%, depending on ablation modality, lesion set complexity, and patient risk profile. Differences in reported incidence proportions were further influenced by variability in endpoint definitions and event-capture windows across trials, including intraprocedural, ≤7-day, and extended follow-up periods.

#### Vascular Access Complications

The pooled incidence of vascular access complications was 0.01 (95% CI, 0.00–0.01) (Figure 3). These events included hematoma, bleeding requiring transfusion, pseudoaneurysm, and arteriovenous fistula as defined in the source trials. Stratified estimates by ablation strategy (PVI vs. PVI+) and energy modality (RF, CRYO, PF) are shown in Figure S4-S5.

**Figure 3.**
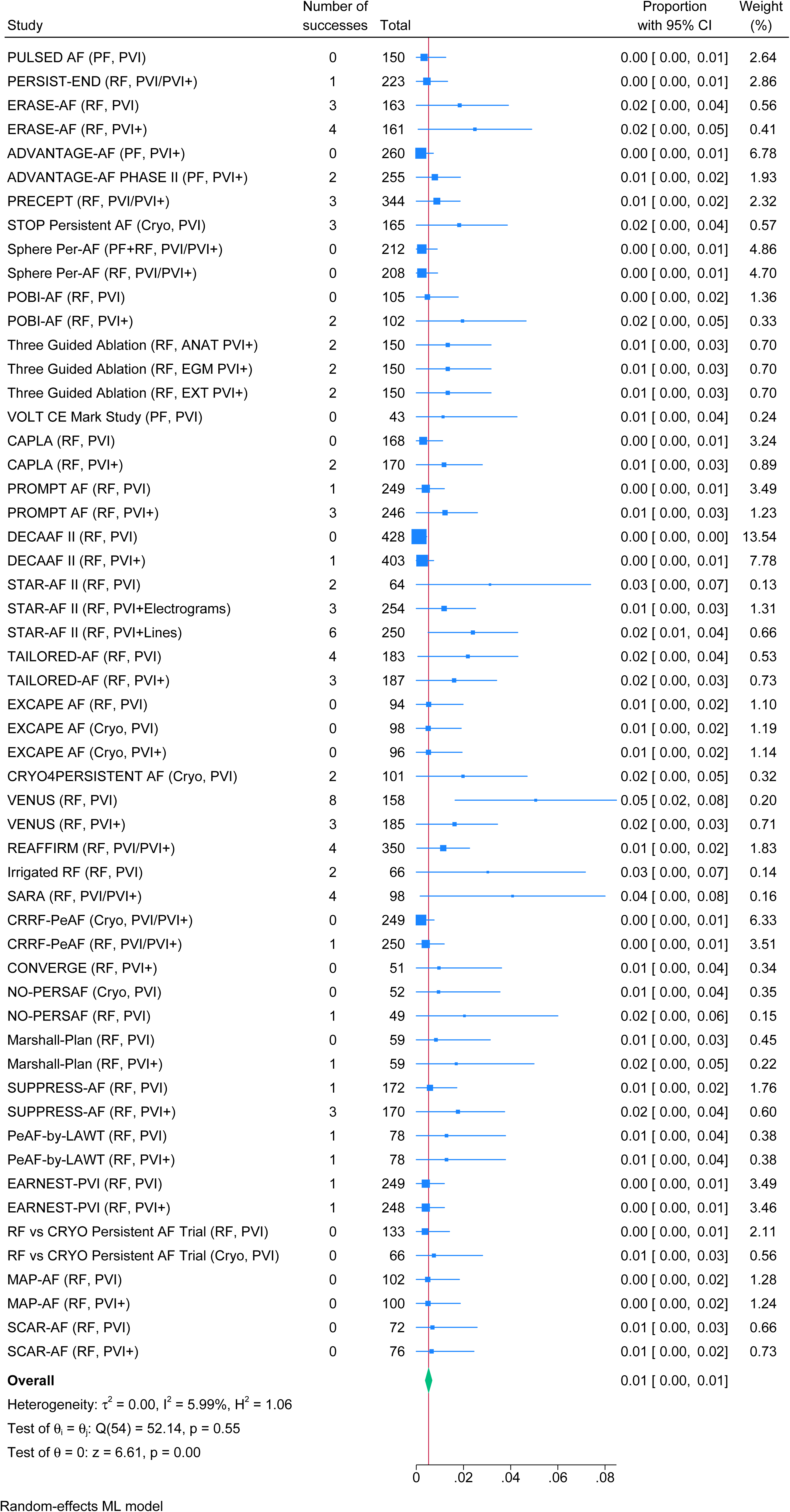
Overall vascular access complications following catheter ablation for persistent atrial fibrillation. CI = confidence interval; proportions represent incidence of vascular access complication per study; squares indicate study estimates (size proportional to weight) with 95% CIs; diamond indicates pooled estimate; random-effects REML model used; heterogeneity assessed by τ², I², and H².

#### Pericardial Complications

The pooled incidence of pericardial complications was 0.01 (95% CI, 0.01–0.01) (Figure 4). This category encompassed pericardial effusion and cardiac tamponade as reported by the individual trials. Stratified estimates by ablation strategy (PVI vs. PVI+) and energy modality (RF, CRYO, PF) are shown in Figure S6-S7.

**Figure 4.**
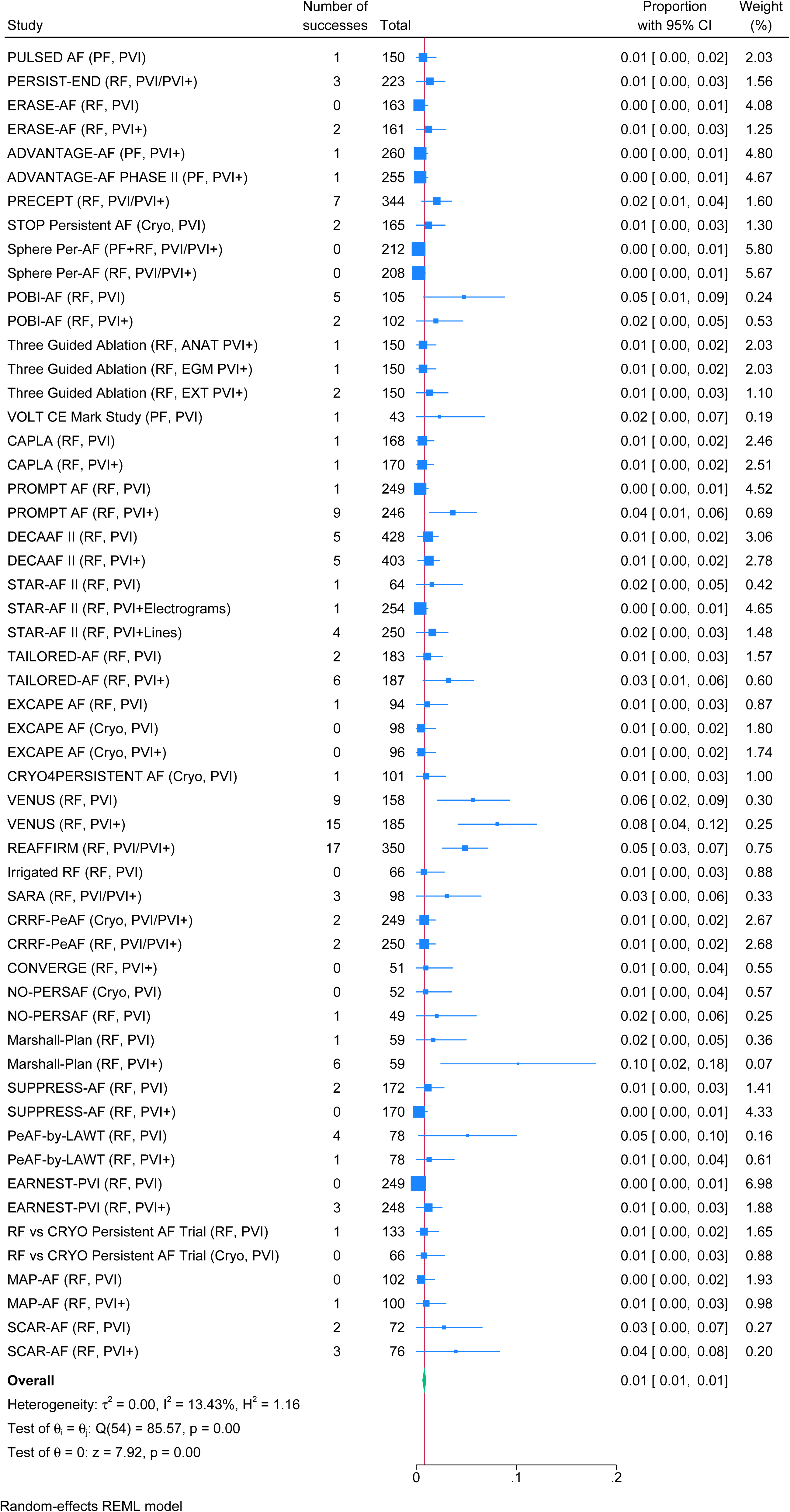
Overall pericardial complications following catheter ablation for persistent atrial fibrillation. CI = confidence interval; proportions represent incidence of pericardial complication per study; squares indicate study estimates (size proportional to weight) with 95% CIs; diamond indicates pooled estimate; random-effects REML model used; heterogeneity assessed by τ², I², and H².

#### Thromboembolic Events

The pooled incidence of thromboembolic events was 0.00 (95% CI, 0.00–0.00) (Figure S8). These events included stroke and transient ischemic attack within the 12-month follow-up window. Stratified estimates by ablation strategy (PVI vs. PVI+) and energy modality (RF, CRYO, PF) are shown in Figure S9-S10.

#### Accessory Organ Injuries

The pooled incidence of accessory organ injuries was 0.00 (95% CI, 0.00–0.00) (Figure S11). This category included atrioesophageal fistula, phrenic nerve injury, and pulmonary vein stenosis as defined in the source publications. Stratified estimates by ablation strategy (PVI vs. PVI+) and energy modality (RF, CRYO, PF) are shown in Figure S12-S13.

#### All-Cause Mortality

The pooled incidence of all-cause mortality within 12 months of the index procedure was 0.00 (95% CI, 0.00–0.00) (Figure S14). Stratified estimates by ablation strategy (PVI vs. PVI+) and energy modality (RF, CRYO, PF) are shown in Figure S15-S16.

### RISK OF BIAS ASSESSMENT

To evaluate potential bias in the included RCTs, the RoB tool was applied across five domains to determine the overall risk for each study. All trials were classified as low risk in the domains of deviations from intended interventions, missing outcome data, and selection of the reported result. While most trials demonstrated low to moderate risk across domains, a high RoB was identified in the measurement of outcomes domain. Two independent reviewers (YC and AH) assessed each trial, with no discrepancies remaining after discussion. Results are shown in Figure S17.

### PUBLICATION BIAS ASSESSMENT

A funnel plot was generated to assess study precision and potential publication bias. Visual inspection revealed no clear asymmetry, suggesting no strong evidence of publication bias and supporting the robustness of the meta-analysis findings (Figure S18).

## DISCUSSION

This meta-analysis of contemporary clinical trials provides an updated view of CA for PsAF. Across randomized and prospective single-arm studies, the pooled 12-month arrhythmia-free proportion was about 65%, with generally low incidence of reported adverse events, reflecting outcomes in a rapidly evolving ablation landscape.

### EFFECTIVENESS OF CONTEMPORARY ABLATION

Compared with earlier meta-analyses of cohorts treated mainly from 2001 to 2015, pooled effectiveness appears modestly higher, likely reflecting advances in catheter technology, lesion delivery, and procedural standardization [42,43]. Reported 12-month success still varied widely (about 38%–88%), consistent with prior reviews showing substantial heterogeneity due to differences in outcome definitions, rhythm monitoring, ablation strategies, lesion sets, patient populations, and trial design [44].

In prespecified subgroup analyses, pooled arrhythmia-free proportions were about 7% higher in studies using recurrence-only endpoints than in those using composite endpoints, with overlapping 95% CIs. This suggests endpoint definition had only a modest effect on pooled estimates, while variability was more likely driven by patient selection, ablation strategy, and follow-up methods.

Overall, the pooled estimate suggests durable rhythm control is achievable in many patients with PsAF. Importantly, these estimates largely reflect protocol-defined single-procedure outcomes and therefore may understate real-world long-term success, where repeat ablation and adjunctive antiarrhythmic therapy are commonly used to enhance arrhythmia-free survival beyond the initial treatment [42,43].

#### Differing Ablation Strategies: Incremental Benefit and Ongoing Uncertainty

Across randomized and single-arm studies, PVI+ strategies were associated with numerically higher freedom from atrial arrhythmia than PVI alone, by about 3%, although effect sizes varied and CIs often overlapped. These findings should be interpreted descriptively rather than as comparative effectiveness because pooled relative effect measures were not calculated and between-study heterogeneity was substantial. Several randomized trials, including ERASE-AF [15], Three-Guided Ablation [22], and TAILORED-AF [11], reported better arrhythmia-free survival with adjunctive or guided ablation beyond PVI alone, whereas CAPLA [27], POBI-AF [20], and PROMPT-AF [21] found no significant incremental benefit from PWI or additional lesion sets.

The broader PsAF literature remains mixed. Some studies show comparable outcomes with PVI alone, and the REAFFIRM trial [29] showed no benefit from added strategies such as focal impulse and rotor modulation (FIRM)-guided ablation, whereas others report benefit from adjunctive left atrial ablation beyond PVI in selected patients [45]. These differences likely reflect atrial substrate, disease chronicity, lesion durability, procedural execution, and operator experience rather than a simple PVI-versus-PVI+ distinction. More evidence is needed to identify which patients benefit from adjunctive ablation and to support a personalized, substrate-guided approach [46].

#### Ablation Energy and Introduction of Pulsed Field Ablation

By energy source, PF and RF ablation showed similar pooled effectiveness and were numerically higher than CRYO, though differences should be interpreted descriptively given overlapping CIs, early-phase designs, and limited follow-up. The PF evidence base in PsAF remains small and largely early phase or single arm, which may overestimate effectiveness. Still, the observed effectiveness of PF ablation in PsAF is encouraging, particularly in the context of its favorable safety profile and the consistency of lesion delivery afforded by myocardial tissue selectivity [47]. Ongoing randomized trials with longer follow-up are needed to determine whether any apparent advantage persists and whether it reflects the energy source itself or differences in lesion-set design and procedural strategy.

### ADVERSE EVENTS ACROSS STRATEGIES AND TECHNOLOGIES

Adverse event incidence proportions across included trials were low overall but heterogeneous, ranging from 0% to about 15%. Vascular access and pericardial complications each occurred in about 1%, while thromboembolic events, accessory organ injuries, and mortality were rare, supporting the favorable safety profile of contemporary PsAF ablation.

Subgroup analyses showed modest differences across strategies and modalities. Pericardial complications approached 0% with PF versus about 1% with RF and CRYO, while vascular access complications were more common with RF and PVI+ strategies, likely reflecting procedural complexity and lesion delivery. Thermal ablation has historically been associated with a higher risk of certain collateral effects, although this relationship is multifactorial and operator dependent. Despite these differences, event incidence proportions were broadly comparable between PVI-only and PVI+ strategies, and overlapping 95% CIs suggest adjunctive lesion sets do not add substantial procedural burden at a population level when performed in experienced centers.

The low complication incidences observed with PF ablation are notable and align with early reports suggesting that its nonthermal, myocardial-selective mechanism may reduce collateral injury, including esophageal and phrenic nerve injury. However, emerging real-world data also show distinct patterns of myocardial injury, hemolysis, and transient renal dysfunction that remain incompletely characterized [48]. These findings should therefore be interpreted cautiously given heterogeneous definitions, variable reporting windows, and limited long-term follow-up, and warrant continued standardized reporting and post-approval surveillance.

### HETEROGENEITY AND INTERPRETATION OF POOLED ESTIMATES

Substantial between-study heterogeneity was observed for both effectiveness and adverse-event outcomes, reflecting differences in patient populations, ablation strategies, lesion sets, rhythm monitoring intensity, endpoint definitions, and trial design. Accordingly, pooled estimates should be interpreted as summary measures across heterogeneous studies rather than precise estimates for any single strategy or population.

A major source of heterogeneity was post-ablation rhythm monitoring. More intensive or continuous monitoring likely applied stricter detection thresholds and may partly explain the wide range of reported 12-month arrhythmia-free proportions, as post-ablation atrial tachyarrhythmia recurrence depends strongly on monitoring strategy [49], and between-trial differences may reflect monitoring protocols rather than true differences in ablation efficacy. Differences in monitoring methods should therefore be considered when interpreting pooled effectiveness estimates or comparing outcomes across strategies and technologies.

### COMPARISON WITH PRIOR LITERATURE

Compared with earlier meta-analyses of PsAF ablation [42,43], this study reflects a more contemporary evidence base, including guided ablation strategies and next-generation technologies. Single-procedure effectiveness has also improved over time, suggesting that better outcomes relate less to lesion extensiveness than to advances in strategy, energy modality, patient selection, mechanistic targeting, and lesion durability. These findings reinforce CA as an effective rhythm-control strategy for PsAF and highlight the need to further optimize technologies, lesion delivery, and patient-specific procedural approaches.

### LIMITATIONS

Several limitations should be acknowledged. First, although a comprehensive literature search was performed, it is possible that not all relevant studies were identified [5]. Second, to maintain consistency in study design and endpoint assessment, we excluded observational studies and registries; thus, findings are limited to clinical trials and may not fully reflect real-world practice. Third, pooling single-arm studies with RCTs may reduce interpretability of the pooled estimates, as RCTs uniquely control for confounding through randomized between-arm comparisons, which are not preserved when all studies are combined into a single absolute-effect summary. Fourth, effectiveness estimates were based on the most detailed data available; when absolute event counts were unavailable, outcomes were derived from published Kaplan–Meier curves or time-specific rates, introducing additional uncertainty [50]. Fifth, adverse event outcomes should be interpreted descriptively because of variability in primary safety endpoint definitions, monitoring strategies, and reporting windows across studies, as noted in contemporary AF ablation consensus guidelines [44]. Sixth, effectiveness endpoint definitions also differed, with some trials reporting arrhythmia recurrence alone and others incorporating repeat ablation or therapy escalation, which may contribute to residual heterogeneity.

## CONCLUSIONS/IMPLICATIONS

This contemporary meta-analysis demonstrates that CA for PsAF is associated with moderate effectiveness and low overall complication incidences in modern clinical trials. While PVI+ strategies and newer technologies such as PF ablation show promise, no single approach consistently outperforms others across all patient populations. These findings support a tailored approach to PsAF ablation that emphasizes patient selection, mechanistic targeting, and procedural durability rather than uniform escalation of lesion complexity.

Future progress in PsAF ablation will likely depend on prospectively designed, adequately powered randomized trials with standardized lesion sets, rigorous and uniform rhythm monitoring, and transparent safety reporting. These efforts are essential to refine ablation strategies, define the role of emerging technologies, and ultimately improve long-term outcomes for patients with PsAF.

## Data Availability

All data analyzed in this study were extracted from previously published clinical trials and are available within the article and its supplemental tables. No new primary data were generated.

## ACKNOWLEDGMENTS

Arshan Harizavi (Conceptualization, Data curation, Methodology, Formal analysis, Manuscript drafting, Visualization, Manuscript review and editing), Yan Chai (Conceptualization, Data curation, Methodology, Meta-analysis support, Manuscript review and editing), Jiayuan Wang (Data curation, Manuscript review and editing), Tiffany Tan (Conceptualization, Methodology, Manuscript review and editing, Supervision).

## NON-STANDARD ABBREVIATIONS AND ACRONYMS

AAD: antiarrhythmic drug
AF: atrial fibrillation
AMSTAR: A MeaSurement Tool to Assess Systematic Reviews
CA: catheter ablation
CI: confidence interval
CRYO: cryoballoon
ECG: intermittent electrocardiogram
EIVOM: vein of Marshall ethanol infusion
FIRM: focal impulse and rotor modulation
ICM: implantable cardiac monitor
LsPAF: long-standing persistent atrial fibrillation
PAF: paroxysmal atrial fibrillation
PF: pulsed field
PRISMA: Preferred Reporting Items for Systematic Reviews and Meta-Analyses
PsAF: persistent atrial fibrillation
PVI: pulmonary vein isolation
PVI+: pulmonary vein isolation with adjunctive lesion sets
PWI: posterior wall isolation
RCT: randomized controlled trial
RF: radiofrequency
RoB: Risk of Bias
SM: substrate modification

